# Single-Cell Nanomotion and Machine Learning for Parallel Bacterial Identification and Antibiotic Screening

**DOI:** 10.1101/2025.11.26.25340999

**Authors:** Santiago Mendoza-Silva, Farbod Alijani, Le-Vaughn Naarden, Roxan Broer, Leo Smeets, Tabea Riepe, Irek Roslon, Aleksandre Japaridze

**Affiliations:** Department of Precision and Microsystem Engineering, Delft University of Technology, Delft 2628 CD, The Netherlands; SoundCell B.V., Molengraaffsingel 12, 2629 JD Delft, The Netherlands; Reinier Haga MDC, Reinier de Graafweg 7, 2625 AD Delft, The Netherlands

**Keywords:** Nanomotion Detection, Graphene Biosensors, Machine Learning, Bacterial Identification, Antibiotic Susceptibility Testing

## Abstract

Rapid and accurate identification of bacterial infections and their resistance to antibiotics is critical to effective clinical decision-making and combating antimicrobial resistance. However, current diagnostic approaches are typically segmented: techniques such as MALDI-TOF provide species identification, but cannot assess antibiotic susceptibility, while standard antimicrobial susceptibility (AST) tests are time-consuming and lack concurrent identification capability. In this study, we overcome these limitations by integrating single-cell nanomotion detection using graphene drums with machine learning (ML) algorithms to perform both tasks simultaneously within a single measurement. Nanomotion signals, nanoscale vibrations from single living cells, are recorded in real-time and transformed into time-frequency spectrograms, which serve as inputs to ML models trained for robust pattern recognition. Our framework enables the differentiation of *Escherichia coli, Staphylococcus aureus*, and *Klebsiella pneumoniae*, while simultaneously distinguishing resistant and susceptible strains with 98% precision. By coupling highly-sensitive graphene nanomotion sensors with advanced ML tools, our approach delivers a label-free bacterial diagnostics, offering both identification and susceptibility profiling at the single-cell level within a couple of hours.

## 1 Introduction

Antimicrobial resistance (AMR) has become one of the most pressing public health threats on a global scale. Drug-resistant infections are currently responsible for an estimated 1.3 million direct deaths worldwide, while claiming more than 35,000 lives annually in the European Union (EU) alone^[1;2]^. Rapid diagnostics are needed to combat AMR^[3]^, with simultaneous pathogen identification and antimicrobial susceptibility testing (AST).

Accurate bacterial identification is the first step in clinical microbiology to direct suitable treatment and steer infection control^[4]^. Efficient diagnosis is crucial for reducing morbidity, mortality, and associated costs. While accurate identification can prevent unnecessary antibiotic use, the prevalent culture-based methods prove laborious, often requiring days to pinpoint pathogen species and longer to generate antibiotic susceptibility profiles^[5]^. This latency often prompts physicians to prescribe broad-spectrum antibiotics, in conjunction with these culture-based methods that typically require 16 to 96 hours until a correct prescription is found^[6]^.

Current identification methods span culture-based methods that grow microorganisms on selective media, molecular (nucleic acid-based) methods like PCR to detect genetic material, and immunological methods that identify pathogen-specific antigens or antibodies^[7]^. More advanced approaches like the Matrix-Assisted Laser Desorption/Ionization Time-of-Flight (MALDI-TOF) mass spectrometry has notably revolutionized both the identification and characterization of bacterial species. By measuring the mass-to-charge ratio of ionized molecules in a sample, MALDI-TOF not only enables the detection of unique bacterial signatures but also facilitates the identification of enzymes that bacteria produce to inactivate antibiotic molecules^[8]^. However, MALDI-TOF is sensitive to experimental conditions and its spectra can vary due to growth conditions and extraction methods, highlighting the need for alternative procedures^[4;9;10;11]^. Moreover, while specific MALDI-TOF variations can detect antibiotic resistance, they fail to provide the minimum inhibitory concentration (MIC) of the antibiotic needed to direct appropriate treatment strategies.

In contemporary medical microbiology AST is the golden standard tool to evaluate how bacterial strains respond to various antibiotics. Methods such as broth microdilution, Kirby-Bauer disk diffusion, E-test, or automated techniques such as VITEK 2 (BioMerieux) and Phoenix (BD) are effective procedures developed to evaluate bacterial susceptibility to antibiotics. Unfortunately, these methods are slow and display variable efficacy based on the bacterial concentrations in the samples. Importantly, no current method offers simultaneous rapid strain identification and AST^[5]^, highlighting the need for advanced techniques.

Over the past decade, nanomotion detection has emerged as a powerful technique for rapid AST^[12;13;14]^. By monitoring nanoscale oscillations generated by living microorganisms, this approach enables growth-independent assessment of bacterial activity, offering a rapid and label-free alternative to conventional culture-based methods. Although nanomotion spectroscopy exhibits considerable promise, its potential to simultaneously conduct bacterial identification and AST remains a challenge. This is because existing nanomotion signals obtained from AFM cantilevers typically represent averaged motions emanating from populations of hundreds of individual bacteria^[15;16;13;14;17]^. Such aggregate signals make precise bacterial identification challenging, as individual bacterial vibrational profiles are crucial for accurate species differentiation^[18]^. To overcome these limitations, recent advances have culminated in the use of graphene drums as nanomoton sensors, pushing the detection sensitivity to single cells^[19;20;21]^. This development potentially provides a renewed opportunity to explore the capability of nanomotion spectroscopy for simultaneous bacterial identification and AST.

In this work, we explore the potential of using single-cell nanomotion signals from graphene drums for simultaneous bacterial identification and rapid AST. Our study probes the efficacy of Machine Learning algorithms such as Convolutional Neural Networks (CNNs) and Support Vector Machines (SVM), in discriminating between three different species of clinically relevant pathogens, including *Escherichia coli, Staphylococcus aureus*, and *Klebsiella pneumoniae*. In addition, these algorithms are adeptly selected to distinguish not only between bacterial species but also between antibiotic-resistant and susceptible strains. Within this framework, a total of 456 data measurements were utilized for the species identification study, and a total of 347 data measurements were employed for the antibiotic susceptibility study. Bacterial nanomotion signals were transformed into time-frequency spectrograms, serving as inputs for these ML algorithms. With an emphasis on pattern recognition, the algorithms were utilized to identify intricate features within the spectrogram images, culminating in the construction of robust classification models. In addition, the performance of these models was critically evaluated using established metrics in ML theory such as accuracy, sensitivity, specificity, and area under the Receiver Operating Characteristic (ROC) curve, while considering computational efficiency. This study pioneers the integration of ML algorithms with single-cell nanomotion detection, targeting bacterial species identification and AST. Our data shows that we can distinguish between *Escherichia coli, Staphylococcus aureus*, and *Klebsiella pneumoniae* species while differentiating resistant from susceptible strains with high precision. By integrating ultra-sensitive graphene nanomotion sensors with advanced machine learning tools, our approach provides a paradigm shift in diagnostics, delivering both rapid species identification and susceptibility profiling in parallel within just a few hours.

## 2 Results

### 2.1 Nanomotion Measurement and Signal Processing

Our experiments were performed on bi-layer CVD graphene, transferred over circular cavities with dimensions of 8 µm in diameter and 285 nm in depth, etched into SiO_2_ as previously described^[19;20]^. Bacteria were first grown overnight on LB-Agar medium at 30°. The next day, a small amount (1:100 volume ratio) of this culture was transferred into fresh LB medium and grown for another 2.5 hours at 30°, until the cells reached an optical density (OD) of about 0.2. This bacterial suspension was then added to the fluidic chamber and left for 10 minutes so the bacteria could settle on the graphene surface. To test how the bacteria respond to antibiotics, clinical isolates of *E. coli* cells were treated with 8 µg/mL of meropenem for one hour. The antibiotic, dissolved at the final concentration in LB medium, was flowed directly into the chamber, after which new measurements were taken on the same graphene sensor array. The chamber was then placed in an interferometric setup, where bacterial nanomotion was measured. The resulting deflections of each graphene membrane due to intrinsic nanomotion of individual microorganisms were read-out optically using a red laser (*λ*=633 nm) (see Figure 1). The reflected signal from the drum was captured by a photodiode and recorded using an oscilloscope (See Methods Section 4.1). The resulting timetraces were used to build spectrograms that formed the basis for ML algorithms. Analysis of these signals revealed that the nanomotion induced by individual microorganisms manifested as random oscillations of the graphene drum, consistent with our earlier observations^[19;20]^.

**Figure 1:**
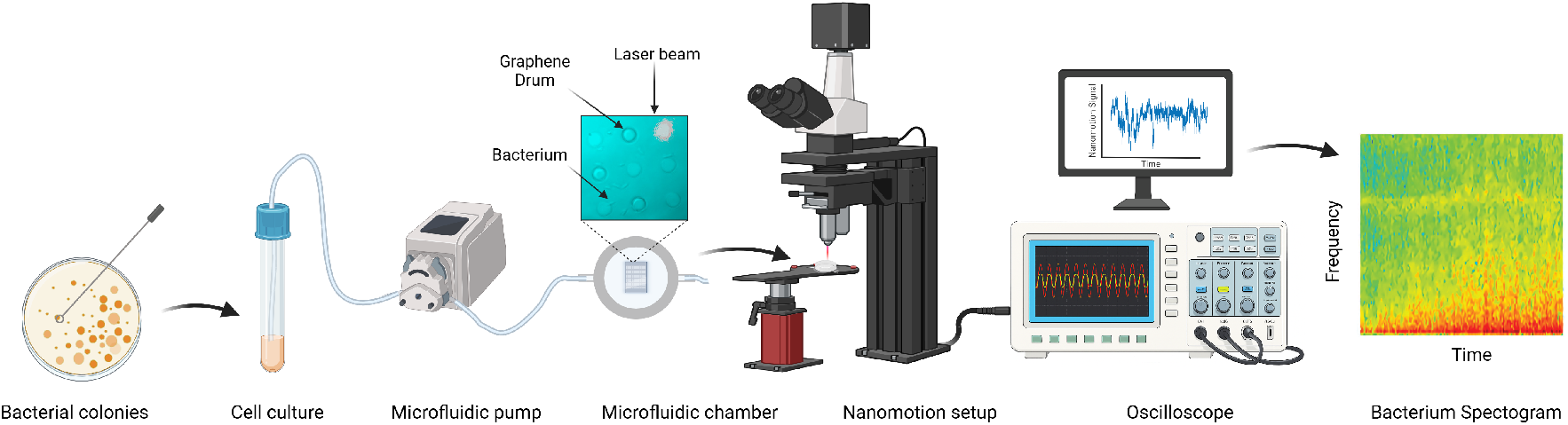
Illustration of the summary of the methodology for bacterial nanomotion measurement to bacterial spectogram generation.Schematic illustration of the procedure: picking up single colonies of bacteria from LB-agar plates, resuspending them in LB growth broth, using microfluidic system to pump live cells into the fluidic chamber where the graphene chips are mounted (inset shows optical image of graphene drums with live bacteria attached and a focused laser beam), followed by the interferometric setup for capturing nanomotion signals, through to the employment of the Short-Time Fourier Transform for generating the spectrograms based on single nanomotion signals.

Measurements were acquired over 30-second intervals and each was annotated with the corresponding bacterial strain and experimental conditions (see Table 1), forming the basis of the dataset used for classification using ML. Since nanomotion signals lack periodicity, analyzing the time-domain data alone does not adequately represent their most relevant characteristics. Therefore, we transformed the signals into a representation that preserves both temporal and spectral information. Consequently, the time-domain signals were segmented using a window function and converted to the frequency domain using a Short-Time Fourier Transform (STFT), yielding typical spectrograms such as the one in Figure 1. These spectrograms can be interpreted as images or maps, which provide a more informative structure for subsequent analysis and enable the use of advanced image-recognition tools. This approach has been shown to outperform methods based solely on frequency domain^[22]^. The resulting spectrograms were then used to train and test ML algorithms for bacterial typing and antibiotic screening as presented in Figure 2. More specifically, trained models were tested to distinguish between different bacterial species and determine antibiotic susceptibility, exclusively based on their nanomotion signatures.

**Table 1:** List of bacterial strains used in the training of both machine learning algorithms.

| Strain List |  |  |  |
| --- | --- | --- | --- |
| *E. coli* | *K. pneumoniae* | *S. aureus* | *E. coli* + 8 $\mu\text{g/mL}$ Meropenem |
| 7740 | KPC2 | L4443 | 7740 (Sensitive) |
|  |  |  | 881 bla-OXA-48 (Sensitive) |
| ATCC 25922 | ATCC 700603 | L4565 | 880 bla-OXA-244 (Sensitive) |
| 1544 bla-OXA-48 |  |  | 1544 bla-OXA-48 (Sensitive) |
| 2139 NDM-5 |  |  | 2139 NDM-5 (Resistant) |
| 1303 NDM-7 |  |  | 1303 NDM-7 (Resistant) |
| 1941 NDM-7 |  |  | 1941 NDM-7 (Resistant) |

**Figure 2:**
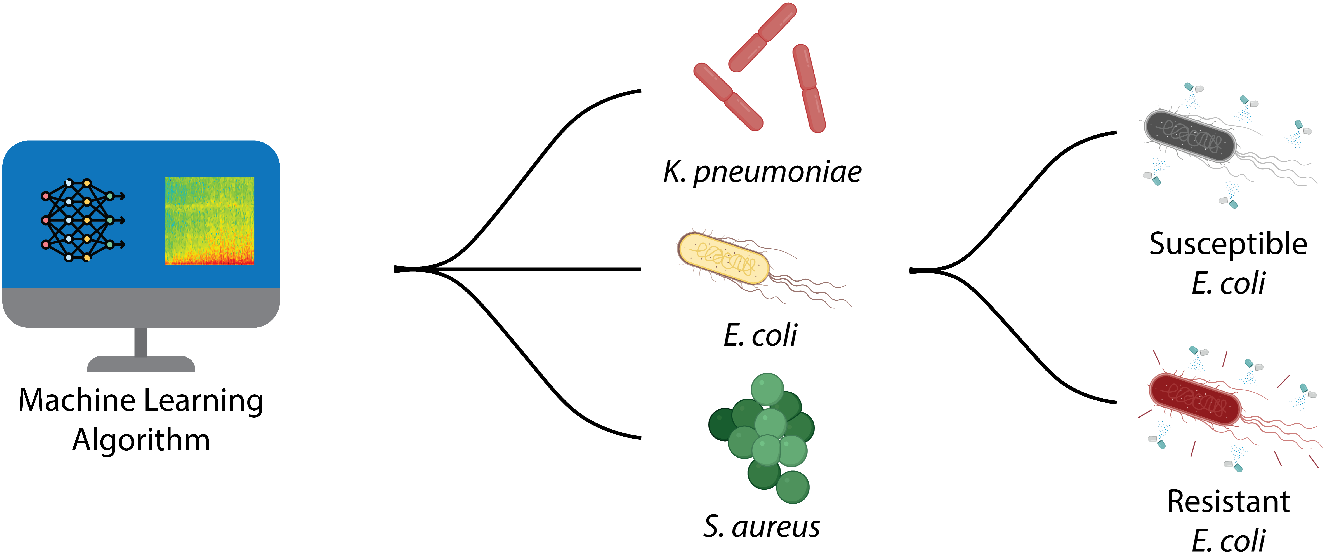
Illustration of the procedure steps, from the input of spectrograms into the ML algorithms for bacterial type identification to discerning susceptible from resistant strains of *E. coli* bacteria.

Figure 3 displays representative spectrograms for different measurement conditions performed in this study. To explore the distinctive signal behavior of microorganisms and assess the applicability of ML for signal classification, we investigated a selection of most common bacterial pathogens, spanning both gram-negative and gram-positive categories, as well as motile and non-motile variants. To generate the associated spectrograms, the time-domain nanomotion signals were segmented using a window of 0.256 s with 50% overlap and converted into the frequency domain using STFT. These parameters were chosen to capture essential spectral features while minimizing artifacts such as spectral leakage. To ensure consistency across all spectrograms, a power spectral density (PSD) scale was used, with endpoints determined by the average maxima and minima over the full dataset.

**Figure 3:**
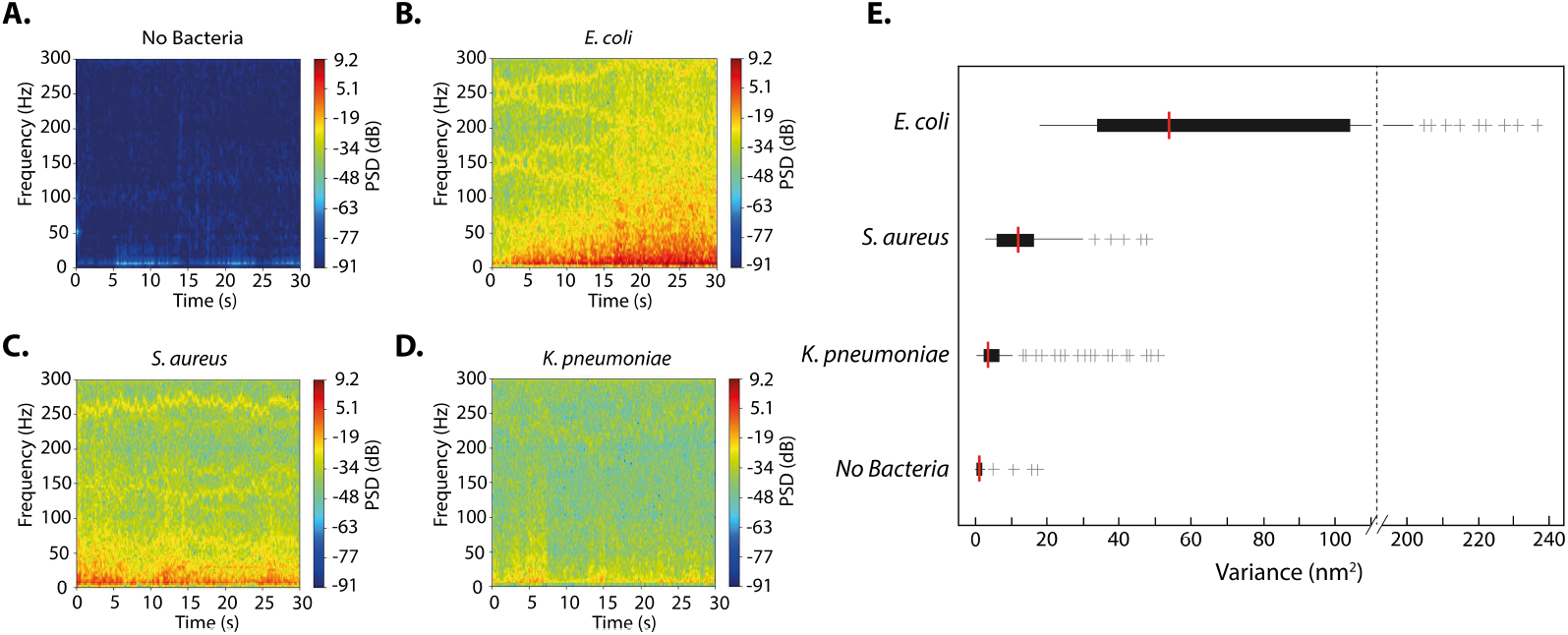
Visualization of single-cell bacterial spectrograms and their respective variances. **A)** Spectrogram for a single empty drum (without bacteria). **B)** Spectrogram for a single *E. coli* bacterium. **C)** Spectrogram for a single *S. aureus* bacterium. **D)** Spectrogram for a single *K. pneumoniae* bacterium. **E)** Box plot for all type of bacteria measurements on graphene as well as for empty control cavities without bacteria. The number of individual traces for each species are as follows: N(*E*.*coli*) = 1130, N(graphene)=928, N(*K*.*pneumoniae*)=526, N(*S*.*aureus*)=204. Outliers are marked with crosses.

Figure 3a shows the spectrogram of an empty graphene drum, i.e., one without bacteria, where the absence of cellular activity is evident in the overall features of the spectogram. However, several stationary components are visible as horizontal traces, which originate from electronic noise in the measurement setup. When live cells adhere to the graphene surface, the resulting spectograms display more prominent features. The spectrograms of *E. coli* show broad, intense activity across a wide frequency range, consistent with its motile behavior. In contrast, the non-motile *S. aureus* bacterium (see Figure 3c) exhibits lower PSD and a narrower frequency band, consistent with its non-motile nature. *K. pneumoniae* (see Figure 3d) represents an intermediate case: although also non-motile, its spectrogram still reveals broad pulses, but at lower intensity compared to *E. coli*. These differences highlight the more stationary character of *K. pneumoniae* relative to *E. coli*, while still distinguishing it from *S. aureus*.

The goal of our study was to apply ML models for two complementary tasks: the first, predicting the antibiotic susceptibility profile of clinically relevant bacteria (*E. coli* to the antibiotic meropenem) and second, identifying various species of bacteria (*E*.*coli, S. aureus*, and *K. pneumoniae*) using the nanomotion signal. To achieve each goal, we used machine learning-based classifying algorithms each customized for the specific task. Performance metrics included accuracy, sensitivity, specificity, confusion matrices and the area under the ROC curve, with emphasis on low false positive rates.

### 2.2 AST Results

The first step of the study was to apply ML models designed to distinguish between susceptible and resistant *E. coli* bacteria exposed to the antibiotic meropenem. Meropenem is a broad-spectrum *β*-lactam antibiotic of the carbapenem class^[23]^. It inhibits bacterial cell wall synthesis, leading to cell death^[24]^. Because meropenem is resistant to most *β*-lactamases, it remains effective against a wide range of gram-negative and gram-positive bacteria. Given its effectiveness and broad spectrum of activity, meropenem is typically reserved for severe or hospital-acquired infections caused by multidrug-resistant organisms.

We used two distinct models, Convolutional Neural Networks (CNN) and Support Vector Machines (SVM), to test the accuracy of the models in predicting the susceptibility and resistance of bacteria to meropenem from nanomotion data. We selected clinical isolates of *E. coli* with well characterized resistance mechanisms against carbapenem drugs (see strain Table 1). *E. coli* cells were exposed to meropenem for 1 hour and the respective nanomotion traces were used to train both models.

Figure 4a presents the dataset used for training and testing both methods. The data were divided into separate training and testing subsets. For the CNN case, the model was trained over 30 epochs, and dropout layers were incorporated to reduce the risk of overfitting. Figures 4b and Figure 4c illustrate the learning dynamics of the CNN model. The progressive increase in accuracy across epochs indicates effective extraction of discriminative patterns from the input data, while the corresponding decrease in loss reflects improved generalization and reduced overfitting. Figure 4e shows the final performance of the CNN based on the confusion matrix. The model correctly classified all susceptible *E. coli* isolates and achieved a 95% accuracy rate for resistant isolates. Misclassification occurred in 5% of resistant isolates, which were incorrectly labeled as susceptible. Overall, the CNN model displayed an accuracy of 98.57%.

**Figure 4:**
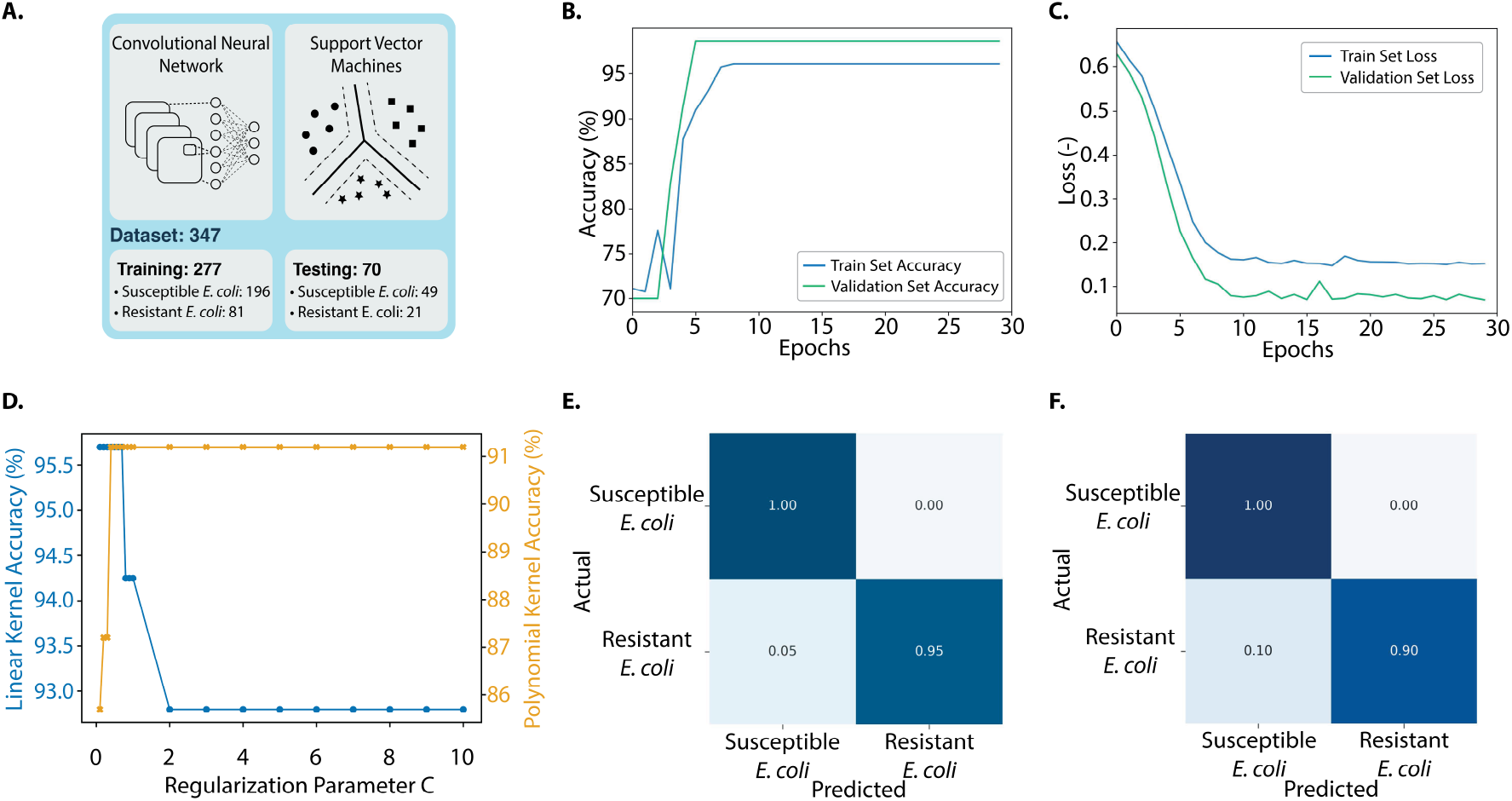
Nanomotion based susceptibility tests using CNN and SVM models to classify resistant and susceptible *E. coli* under antibiotic exposure.**A)** Dataset composition used for training and testing both algorithms. **B)** CNN algorithm training accuracy across epochs. **C)** CNN algorithm Loss value across epochs. **D)** SVM accuracy with linear and 4th-degree polynomial kernels as a function of the regularization parameter *C*. **E)** Confusion matrix of the CNN model showing predictions on the test dataset for classifying resistant and susceptible *E. coli*. The model achieved a total accuracy of 98.57%. **F)** Confusion matrix of the SVM algorithm showing predictions on the test dataset for classifying resistant and susceptible *E. coli*. The model achieved a total accuracy of 97.14%.

Similarly to the CNN model, the SVM algorithm exhibited a notable performance, achieving an overall accuracy of 97.14%. As shown in Figure 4f, the algorithm accurately classified susceptible *E. coli* at a perfect rate of 100% and resistant *E. coli* at 90%. It is interesting to note that the model once again misclassified the resistant *E. coli* as susceptible (at an 10% error rate) indicating that given that the model has more data on susceptible strains it learns more patters from it rather than from resistant strains.

In evaluating the two SVM kernel implementations, both the linear and 4th-degree polynomial kernels achieved peak accuracies close to 96% and 97%, respectively. However, the linear kernel in Figure 4d exhibited greater sensitivity to alterations in the regularization parameter. As the regularization parameter increased, the accuracy decreased significantly from 96% to 89%, indicating a challenge for the model to distinguish unseen data. This suggests that the data in this study might not be linearly separable in the hyperspace where the SVM algorithm is constructing the decision boundary. In contrast, the fourth-degree polynomial kernel in Figure 4c, displayed steadier accuracy rates under similar conditions, reflecting the inherent complex characteristics of the data sets. Given the datasets skewed class distribution and the polynomial kernels consistent performance in capturing non-linear relations to properly separate the data, it emerged as the preferred choice. As a result, SVM with a fourth degree polynomial kernel, using a regularization parameter C = 1.0, was determined to be the optimal selection within the SVM options for this particular study case. Overall, the comparison indicates that while both kernels can achieve high accuracy, the polynomial kernel better identifies an optimal separating hyperplane, offering superior generalization and stability in classifying the spectrograms, and is therefore better suited to the complex nature of the nanomotion data.

### 2.3 Results for Bacteria Species Identification

The next challenge we addressed was using nanomotion signals from single bacteria to determine their species. To tackle this task, we again employed two models, CNN and SVM. Figure 5a shows the total number of samples used to train both methods, considering three different bacterial strains. To enhance the ability of the models to learn relevant spectrogram features and improve their generalization capacity, data augmentation was applied to the training set, introducing random transformations such as rotations, zooming, and shifts. As a result, the accuracy across epochs graph in Figure 5b shows a pronounced rise during the early epochs, indicating efficient pattern recognition from the training data. Although some fluctuations appear throughout training, a common outcome in models employing data augmentation, these likely reflect the variability introduced by the random transformations applied at each epoch. However, the general trend demonstrates a steady increase in accuracy until the training and validation curves converge, most clearly around the 100^th^ epoch. This behavior reflects improved model generalization and a reduced risk of overfitting.

**Figure 5:**
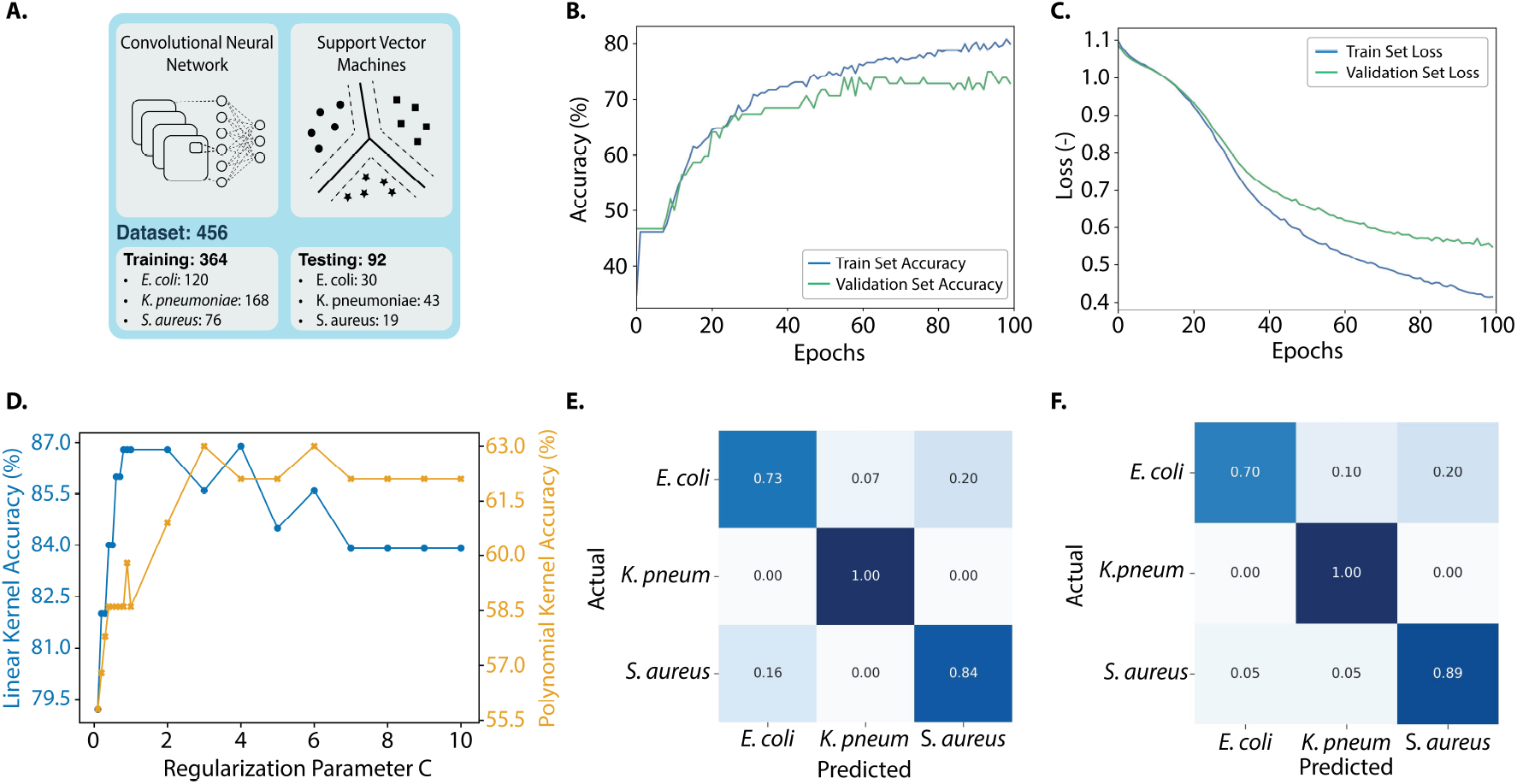
Nanomotion based identification of *E. coli, K. pneumoniae* and *S. aureus* bacteria using CNN and SVM algorithms. **A)** Dataset composition used for training and testing both algorithms. **B)** CNN algorithm training accuracy across epochs. **C)** CNN algorithm Loss value across epochs. **D)** SVM accuracy with linear and 4th-degree polynomial kernels as a function of the regularization parameter *C*. **E)** Confusion matrix of the CNN model showing predictions on the test dataset for bacteria species identification. The model achieved a total accuracy of 88.04%. **F)** Confusion matrix of the SVM algorithm showing predictions on the test dataset for bacteria species identification. The model achieved a total accuracy of 87.04%.

Furthermore, Figure 5c provides additional information about the learning dynamics of the model. The starting loss value of 1.1, resulting from random weight initialization, decreases steadily, showing the proficiency of the model in learning data patterns. The smooth trajectory of the loss function underscores stable learning, suggesting that the hyperparameters, especially the learning rate, are appropriately configured and chosen. Although the training loss remains slightly below the validation loss, indicating mild overfitting, their overall proximity confirms the notable generalization capability of the model, illustrating its adeptness at fitting the training data and generalizing to unseen samples. Consistent with the previous case, the CNN model demonstrated robust performance, achieving an overall accuracy of 88.04%. As shown in the confusion matrix in Figure 5e, the model correctly identified bacterial species with accuracies of 73% for *E. coli*, 100% for *K. pneumoniae*, and 84% for *S. aureus*. Notably, the main inaccuracies of the CNN algorithm occurred in misclassifying *S. aureus* as *E. coli* with an error of 20% and *E. coli* as *S. aureus* at 16%.

In parallel, the SVM algorithm exhibited a significant performance, achieving an overall accuracy of 87.04%. As illustrated in Figure 5f, the species-specific accuracies were recorded as 70% for *E. coli*, 100% for *K. pneumoniae*, and 89% for *S. aureus*. Notable misclassifications included the prediction of *S. aureus* as *E. coli* with a 20% error rate, and *K. pneumoniae* as *E. coli* with a 10% error rate.

Furthermore, for the SVM models used in bacterial differentiation via spectrograms, distinct trends became evident for both the linear and 4th-degree polynomial kernels. The linear kernel, as shown in Figure 5d, achieved peak accuracy at regularization values between 0.8 and 2.0, suggesting an optimal balance between bias and overfitting. However, accuracy declined as values exceeded 4.0, indicating that the spectrogram data are not easily separable with a linear decision boundary with higher values of the regularization parameter *C*. In contrast, the 4th-degree polynomial kernel in Figure 5c, exhibited a steady accuracy increase from 0.1 to 3.0, followed by a plateau between values of 7 to 10, where it maintains a consistent accuracy across varying *C* values, adapting better to the intrinsic characteristics of the dataset at higher values of *C* compared to the linear kernel. These patterns highlight the delicate balance between regularization and SVM generalization when working with the bacteria spectrogram data, which originates from the ability of both to identify an optimal decision hyperplane that accurately separates the data into the different bacterial classes.

During the evaluation of both kernels, peak accuracies of approximately 87% were observed. Notably, the linear kernel, especially with a smaller regularization parameter (*C* = 0.4), demonstrated that the data are largely linearly separable in the transformed feature space at lower *C* values. This suggests that, within this high-dimensional space, the distinct bacterial categories can be separated with relative simplicity. The linear nature of the model not only reduces the risk of overfitting but also enhances interpretability. While the 4th-degree polynomial kernel can capture more complex relationships, its higher complexity increases the risk of overfitting. Given their comparable performance, the linear SVM is favored for its simplicity, robustness, and superior generalization potential. Although the CNN achieved marginally higher overall accuracy, the linear SVM yielded higher accuracy with lower false-positive rates (see Methods 4.3), an essential factor in preventing inappropriate therapeutic interventions and adverse patient outcomes.

## 3 Conclusion

In this study, we demonstrated that single-cell nanomotion signals recorded with graphene nanomotion sensors can be effectively analyzed using ML to perform both bacterial species identification and antibiotic susceptibility testing within a single measurement. By converting nanomotion traces into time–frequency spectrograms and using them as inputs to machine learning algorithms like Convolutional Neural Networks (CNNs) and Support Vector Machines (SVMs), we achieved accurate classification of clinically relevant pathogens and their susceptibility profiles. Our models successfully differentiated *Escherichia coli, Staphylococcus aureus*, and *Klebsiella pneumoniae* species, reaching accuracies of up to 88%, and distinguished resistant from susceptible *E. coli* strains exposed to meropenem with up to 98.6% accuracy. These findings confirm that single-cell nanomotion signals contain discriminative features that capture both species-specific and antibiotic response patterns, which can be efficiently learned through image-based ML approaches, without the need for labeling or culture growth after training.

Both algorithms showed high performance, although their accuracy varied depending on the classification task. For the case of susceptibility profiling, the CNN algorithm achieved the highest performance in detecting antibiotic resistance, reaching 98.6% accuracy for the classification of resistant and susceptible *E. coli* strains, reflecting its ability to extract complex spatial and temporal features from spectrogram representations of nanomotion signals. In contrast, the SVM algorithm performed better for bacterial species identification despite its overall accuracy of 87.04%, as it exhibited a higher generalization across classes when compared to the CNN model. Additional evaluation on false positive rates also corroborated the performance of each method in both classification study cases. Overall, the SVM exhibited more stable generalization across classes, while the CNN captured more detailed spectral variations. These results demonstrate that algorithm selection can be optimized according to diagnostic requirements, balancing classification accuracy and feature complexity.

To sum up, our results show that combining the high sensitivity of graphene nanomotion sensors with ML enables fast, label-free AST and identification of bacteria. Since the trained models analyze nanomotion signals from individual cells, results can be obtained within 1-2 hours, eliminating the need for time-consuming culturing steps. With further development, this approach could establish nanomotion spectroscopy as a powerful platform for real-time diagnostics and for studying cellular biophysics and antimicrobial resistance.

## 4 Methods

### 4.1 Bacterial sample preparation

All bacterial isolates (BSL-2), were anonymous clinical isolates obtained from the medical microbiology department of the Reinier Haga Medical Centre in Delft. Bacteria were grown in LB medium overnight at 30°C to reach the late exponential phase. The next day before performing experiment, the culture was refreshed (1:100 volume) for 2.5 hours on fresh LB medium at 30°C to reach an optical density (OD600) OD= 0.2. The fluidic chamber was filled with the solution at this concentration and left for 10 minutes horizontal position to deposit the bacteria on the graphene surface. The temperature of the fluidic chamber was maintained at 30°C throughout the full measurement. To determine antibiotic susceptibility and resistance, *E. coli* strains were incubated with 8 µg/mL meropenem for 1 hour. Meropenem was dissolved in LB and directly flushed into the liquid chamber and afterwards a new round of measurements was performed on the same array of graphene drums. The chamber was placed in the interferometric setup that was equipped with Attocube ECSx5050 nano positioners that allow automated scanning. The motion of the bacterium caused changes in the optical path, that were monitored by a photodiode and an oscilloscope (Rohde & Schwarz RTB2004). The setup was programmed to run through typically 150-200 measurement points per condition. The measurements were performed in an air-conditioned room with a temperature of 21°C. The substrates were 5 mm × 5 mm silicon chips with a graphene layer. The latter were patterned with circular cavities by a reactive ion etch, where silicon acted as a stop layer, creating cavities with a diameter of 8um, described earlier^[19]^. For species identification and susceptibility determination, the strains as described in Table 1 were used:

### 4.2 Implementation of ML Algorithms

ML algorithms were implemented on spectrograms generated from 30 s signal measurements. The spectrograms were computed using a window size of 0.256 s and an overlap of 0.128 s. A custom Python script, combining SciPy and Matplotlib functions, processed each signal using the spectrogram() function from SciPy to obtain the Power Spectral Density matrix via a Short-Time Fourier Transform (STFT). The nperseg value was set to a power of two for computational efficiency. The resulting spectrograms exhibit continuous and well-resolved time–frequency content.

Next, to perform the classification (see Figure 2), spectrograms obtained from nanomotion signals were used as input for two ML algorithms: Support Vector Machines (SVM) and Convolutional Neural Networks (CNN). SVMs were chosen for their effectiveness in multiclass classification, while CNNs were applied to exploit the image-like structure of spectrograms, automatically learning local features without the need for manual feature engineering^[25]^. Table 2 and Table 3 showcase the architecture used for SVM models while Table 4 and Table 5 showcase the architecture used for CNN algorithms. Figure 6 illustrates the training workflow of both approaches. Each spectrogram was represented as a two-dimensional *n × m* matrix of PSD values from the STFT, where *n* is the number of frequency bins and *m* the number of time windows within the 30-second interval. In our case, *n* = 257 and *m* = 203, resulting in matrices of size 257 × 203, with each entry corresponding to the estimated signal power at a given frequency and time.

**Table 2:** SVM Architecture for the Bacteria Species Identification case.

| Stage | Output Shape | Details | Details |
| --- | --- | --- | --- |
| Input (Train) | (364, 7456) | — | Flattened images |
| Input (Test) | (92, 7456) | — | Flattened images |
| Feature Vector | 7456 | $32 \times 233$ | Grayscale image |
| SVM Classifier | — | Kernel: Linear<br>Degree: 1<br>$C = 0.4$ | scikit-learn SVC<br>Linear kernel<br>Regularization |

**Table 3:** SVM Model Architecture for the Antibiotic Susceptibility Testing case.

| Component | Output Shape | Parameters | Details |
| --- | --- | --- | --- |
| Input (Train) | (276, 7456) | — | Flattened images |
| Input (Test) | (70, 7456) | — | Flattened images |
| Feature Vector | 7456 | $32 \times 233$ | Grayscale image |
| SVM Classifier | — | Kernel: Poly<br>Degree: 4<br>$C = 1.0$ | scikit-learn SVC<br>Polynomial kernel<br>Regularization |

**Table 4:**
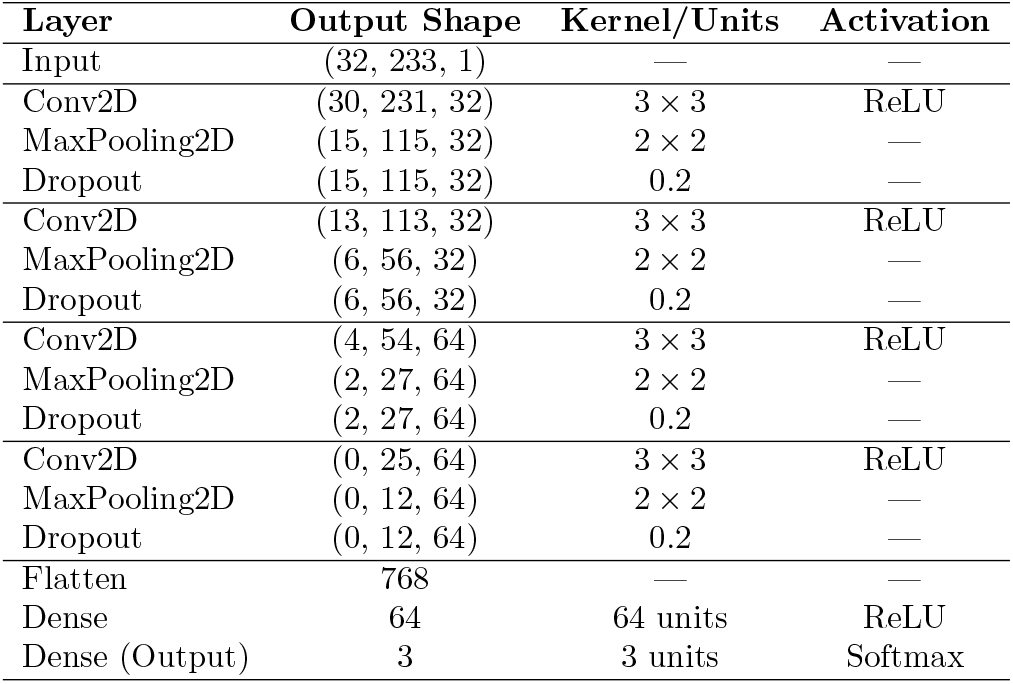
CNN Architecture for the Bacteria Species Identification case.

**Table 5:** CNN Architecture for the Antibiotic Susceptibility Testing case.

| Layer | Output Shape | Kernel/Units | Activation |
| --- | --- | --- | --- |
| Input | (32, 233, 1) | — | — |
| Conv2D | (30, 231, 32) | $3 \times 3$ | ReLU |
| MaxPooling2D | (15, 115, 32) | $2 \times 2$ | — |
| Conv2D | (13, 113, 32) | $3 \times 3$ | ReLU |
| MaxPooling2D | (6, 56, 32) | $2 \times 2$ | — |
| Conv2D | (4, 54, 64) | $3 \times 3$ | ReLU |
| MaxPooling2D | (2, 27, 64) | $2 \times 2$ | — |
| Flatten | 3456 | — | — |
| Dense | 64 | 64 units | ReLU |
| Dense (Output) | 2 | 2 units | Softmax |

**Figure 6:**
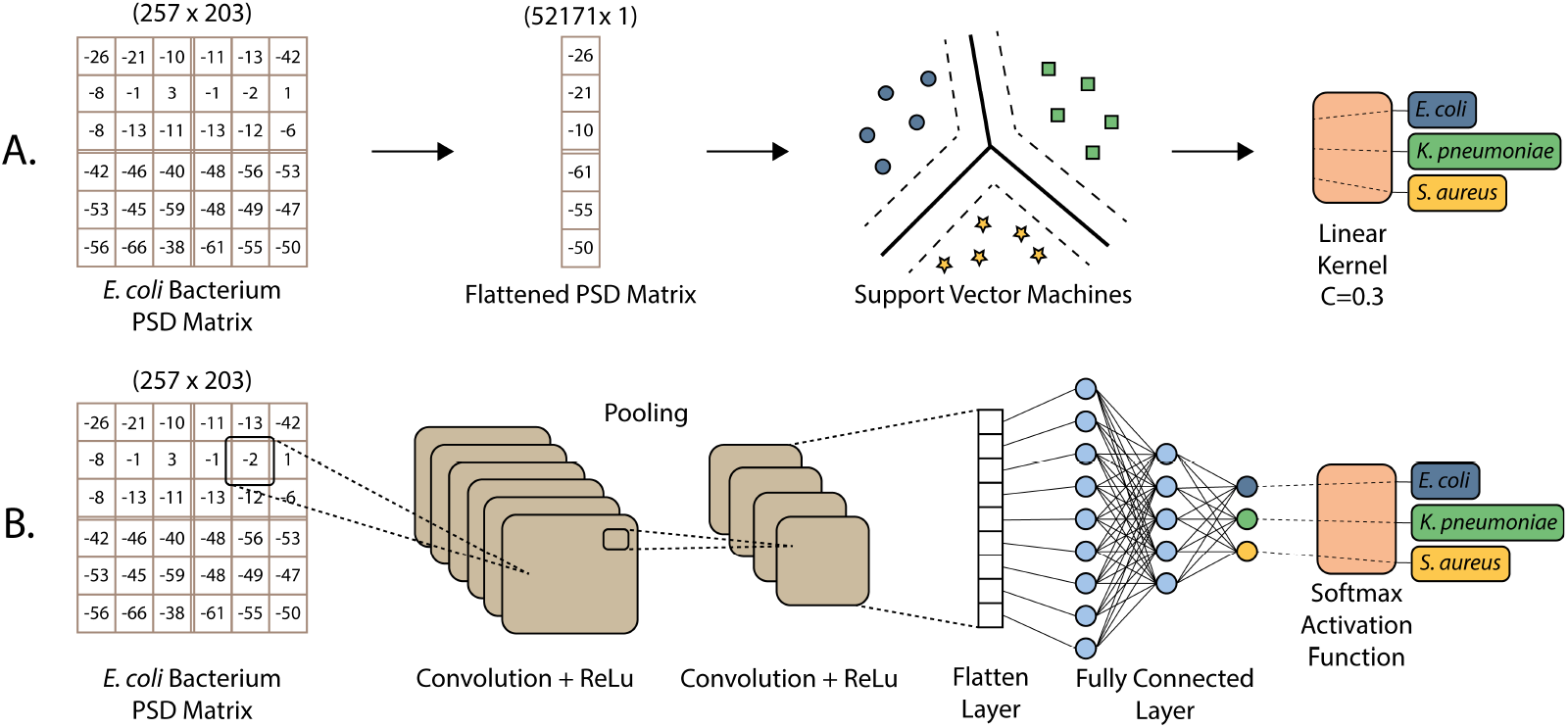
Illustration of the two machine learning algorithms employed in the bacteria species identification study case. **A)** Schematic representation of the applied procedure in the implementation of the SVM algorithm. Spectrograms are employed as Power Spectral Density matrices that are used during training for later performing bacterial classification. **B)** Schematic representation of the applied procedure in the implementation of the CNN algorithm. Power Spectral Density matrices are used in convolution operations to train the algorithm to ultimately perform the bacterial classification task.

For the SVM implementation (Figure 6a, See SI information), the PSD matrices were flattened row by row into one-dimensional feature vectors of length 52,171, producing fixed-length inputs for the classifier. The parameter max iter, which controls the maximum number of iterations for solver converge, was set to its default value of −1, corresponding to an unlimited number of iterations. The SVM then determined optimal separating hyperplanes by maximizing the margin between bacterial categories or resistant versus susceptible cases. To explore kernel effects, both a conventional linear kernel and a polynomial kernel of degree four were tested. In addition, the regularization parameter *C* was used to balance margin maximization against classification errors, penalizing misclassified points to improve generalization. In contrast, the CNN implementation (Figure 6b) used the two-dimensional PSD structure and focused on spatial pattern recognition. Successive convolutional and pooling layers combined with nonlinear activation functions extracted local features and generated high-dimensional feature maps encoding increasingly complex signal representations^[26]^. These feature maps were then flattened and passed to fully connected layers for classification. At the output, a Softmax activation function was employed to produce class probabilities in the multiclass bacterial typing task, while a Sigmoid activation function was used in the antibiotic susceptibility case, where binary classification was required.

### 4.3 Evaluation and Selection of Optimal Algorithmic Approaches

To identify the most suitable algorithm for each classification task, both CNN and SVM models were evaluated, focusing on accuracy, interpretability, and Receiver Operating Characteristic (ROC) analysis. The ROC curve, a standard tool in machine learning, illustrates the trade-off between the True Positive Rate (TPR), reflecting sensitivity, and the False Positive Rate (FPR), reflecting specificity. In this metric, an optimal classifier achieves a high TPR with a low FPR, reflected by an Area Under the Curve (AUC) value close to 1, whereas an AUC of 0.5 indicates random classifier performance. Therefore, curves approaching the top-left corner represent superior model performance.

Figure 7a shows the micro-average ROC curve for species classification using the CNN and linear SVM models. The micro-average ROC was employed because a traditional ROC curve is limited to binary classification, whereas the micro-average provides a comprehensive evaluation in multiclass settings. Although the CNN achieved a slightly higher AUC (0.9469) than the SVM (0.9442), closer inspection of the ROC curves reveals intervals where the SVM outperforms the CNN. These overlaps indicate the ability of SVM to optimize the TPR within specific FPR ranges. In bacterial classification, this distinction is particularly relevant, as minimizing false positives is essential to avoid inappropriate therapeutic interventions that could affect patient outcomes. Given its favorable balance between specificity and sensitivity, the linear SVM emerges as the preferred classifier for bacterial species differentiation, enabling high TPR with minimal FPR.

**Figure 7:**
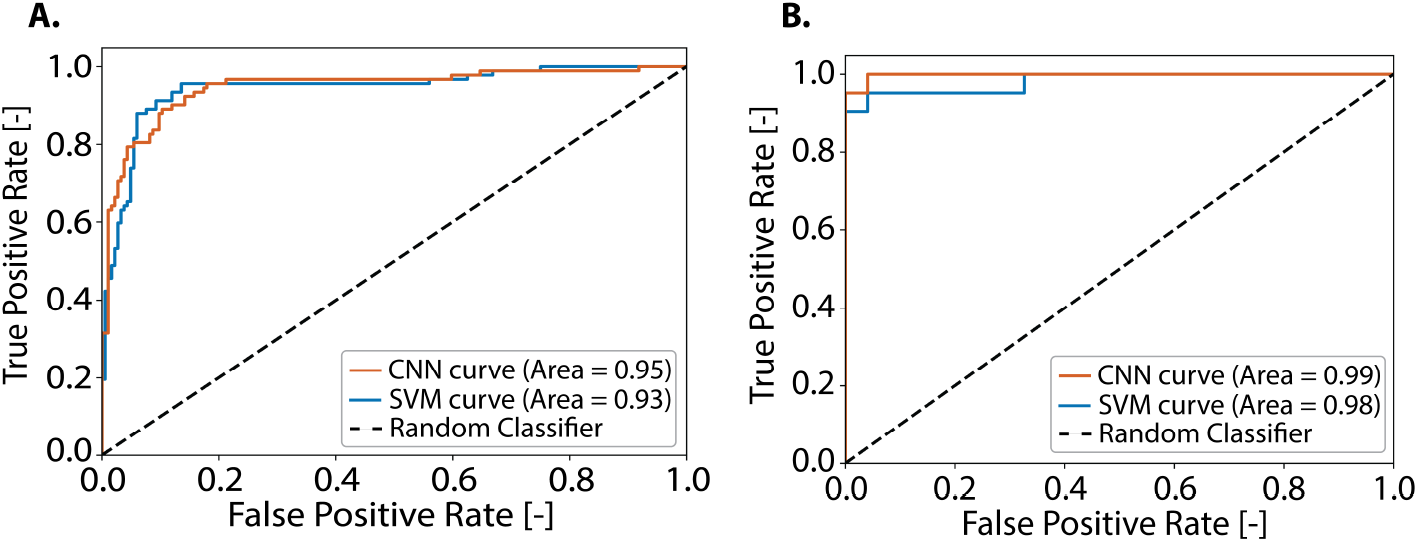
Results for identification of *E. coli, K. pneumoniae* and *S. aureus* bacteria using CNN and SVM algorithms. **A)** Dataset composition used for training and testing both algorithms. **B)** CNN algorithm training accuracy across epochs. **C)** CNN algorithm Loss value across epochs. **D)** SVM accuracy with linear and 4th-degree polynomial kernels as a function of the regularization parameter *C*. **E)** Confusion matrix of the CNN model showing predictions on the test dataset for bacteria species identification. The model achieved a total accuracy of 88.04%. **F)** Confusion matrix of the SVM algorithm showing predictions on the test dataset for bacteria species identification. The model achieved a total accuracy of 87.04%.

In a similar manner, Figure 7b shows the traditional ROC curve for the binary susceptibility classification task using the CNN and 4th polynomial kernel SVM models. The CNN achieved a higher AUC (0.9981) than the 4th-degree polynomial SVM (0.9825). At the critical threshold where false positives begin to rise from zero, the CNN reached a true positive rate of 0.9524, slightly surpassing the SVM’s 0.9048. This difference reflects the CNN’s superior balance between sensitivity and specificity. Given the importance of accurate classification—since even small misclassification rates could lead to incorrect diagnoses—the CNN represents a more reliable approach for distinguishing between susceptible and resistant *E. coli* strains.

## Data availability

The raw datasets of this study are available from the corresponding author on request. The codes used in this study can be found on https://github.com/SantiagoMendozaS/CNN_SVM_bacteria_identification

## Acknowledgments

Financial support was provided from the European Innovation Council Transition grant (no. 10113671) and Kansen voor West Zorgtech Voucher (KVW3-00434). FA further acknowledges financial support from the European Union (ERC Consolidator, NCANTO, 101125458). Views and opinions expressed are however those of the author(s) only and do not necessarily reflect those of the European Union or the European Research Council. Neither the European Union nor the granting authority can be held responsible for them. The authors thank Cees Dekker for fruitful discussions.

## Author Contributions Statement

A.J., and F.A. conceived the idea. S.M. and T. R., performed ML implementation and data analysis with main contribution from S.M. A.J.. LN. and RB. collected the data and performed the experiments.

I.E.R fabricated the graphene sensors. A.J. LS and F.A. designed the experiments. The project was supervised by F.A. and A.J. All authors contributed to the interpretation of the results, writing of the manuscript, with the main contribution from S.M, A.J., and F.A.

## Supplementary Tables

